# Individualized Hybrid Electroconvulsive Therapy (i-HECT) Shows Rapid Anti-Depressant Effect and Improved Cognition in Young Patients with Depression

**DOI:** 10.1101/2024.08.01.24311339

**Authors:** Jing-ya Zhang, Lun Zeng, Jia Li, Mian-mian Chen, Shu-xian Xu, Baijian Tan, Xin-hui Xie

**Affiliations:** Department of Clinical Psychology, Second People’s Hospital of Huizhou, Huizhou, China; Electroconvulsive Therapy Room, Second People’s Hospital of Huizhou, Huizhou, China; Department of Psychiatry, Renmin Hospital of Wuhan University, Wuhan, Hubei, PR China; Brain Function and Psychosomatic Medicine Institute, Second People’s Hospital of Huizhou, Huizhou, China

**Author notes:** Corresponding author: Xin-hui Xie Addresses: 1) Department of Psychiatry, Renmin Hospital of Wuhan University, No. 99 Jiefang Road, Wuchang District, Wuhan, Hubei, PR China. 2) Brain Function and Psychosomatic Medicine Institute, Second People’s Hospital of Huizhou, Huizhou, China. These authors have contributed equally to this work (Jing-ya Zhang, Lun Zeng). Authors’ contributions Jing-ya Zhang: Investigation, Data Curation, Writing - Original Draft; Lun Zeng: Investigation, Data Curation, Writing - Original Draft, Funding acquisition; Jia Li: Investigation, Data Curation; Mian-mian Chen: Visualization, Writing - Review & Editing; Shu-xian Xu: Writing - Review & Editing; Baijian Tan: Supervision, Funding acquisition; Xin-hui Xie: Conceptualization, Methodology, Formal analysis, Writing - Original Draft; Writing - Review & Editing.

**Keywords:** Electroconvulsive Therapy, i-HECT, Hybrid-ECT, rapid antidepressant effect, cognition, safety, side effect

## Abstract

**Background:** For young patients with depression, electroconvulsive therapy (ECT) is highly effective but causes acute cognitive side effects. We designed a new i-HECT therapy combines ECT with low-charge electrotherapy (LCE) and individual symptom monitoring to reduce cognitive impairments.

**Methods:** i-HECT comprised two treatments: ECT and LCE. ECT utilized an energy set of 1.5 times the seizure threshold (ST), while LCE was set at 0.5 ST. The initial session employed ECT. Subsequent sessions involved ECT or LCE, depending on whether meeting the ECT-LCE transition criteria (MADRS total score < 22 or ≥ 50% reduction), assessed after each session.

**Results:** The intention-to-treat analysis revealed an 80.4% response rate and a 58.7% remission rate (Hedges’ g = 3.29). Notably, both subjective and objective cognitive functions significantly improved post-i-HECT treatments and during the 3-month follow-up periods.

**Conclusion:** The i-HECT protocol may provide a rapid antidepressant treatment option with cognitive benefits for young depression patients.

**Highlights:**

- Designed a novel but simple ECT protocol, i-HECT, for young depression patients.
- The trial employed Simon’s optimal two-stage design with a high power of 0.95.
- The i-HECT rapidly improved both depression symptoms and cognitive functions.

## 1. Introduction

Severe depressive symptoms in young patients can result in significant consequences, underscoring the critical need for prompt and efficacious antidepressant interventions. Although electroconvulsive therapy (ECT) exhibits rapid antidepressant efficacy [1], young patients experience acute significant memory and cognitive impairments as side effects [2].

During ECT treatment, symptom reduction is rapid initially, and side effects increase over time. To balance efficacy and side effects, we proposed a new ECT strategy termed Hybrid-ECT. In Hybrid-ECT, the first three sessions use standard bilateral ECT with energy set at 1.5 times the seizure threshold (ST). Thereafter, it transitions to low-charge electrotherapy (LCE) at 0.5 ST. Our previous randomized controlled trial (RCT) demonstrated that Hybrid-ECT maintained similar antidepressant effects to ECT but with significantly fewer side effects [3].

Furthermore, after reviewing data from both Hybrid-ECTs and ECTs, we noted individual variations in the initial response to ECTs. For instance, while some patients experienced a 50% decline in depression scale scores after a single ECT session, others may require more than nine sessions to achieve similar results. The variability suggested the potential for personalized transition conditions from ECT to LCE within Hybrid-ECT, aiming to optimize therapeutic benefits. Therefore, we devised a new but simple protocol called individualized Hybrid-ECT (i-HECT). Under the i-HECT protocol, we assess patients’ symptoms after each ECT session. If the predetermined ECT-LCE transition criteria are met, we implement the switch. To evaluate the antidepressant efficacy and side effects, particularly cognitive, of i-HECT in young patients with depression, we conducted this trial.

## 2. Methods

This single-arm trial employed a Simon’s optimal two-stage design [4] with a 3-month follow-up duration, conducted at the Second People’s Hospital of Huizhou (April 2021–November 2023) in accordance with the Declaration of Helsinki [5]. Both patients and raters remained blinded to the ECT-LCE transition. The Human Ethics Committee of the Hospital approved the protocol. Patients and their legal guardians can withdraw at any time for any reason. Trial Registration: Chinese Clinical Trial Registry (http://www.chictr.org.cn, ChiCTR2100045682).

### 2.1 Patients

Inclusion criteria: 1) Age 16–25 years old; 2) Inpatients diagnosed with unipolar or bipolar depression according to ICD-10, with or without psychotic symptoms; 3) Baseline Montgomery-Åsberg Depression Rating Scale (MADRS) [6] score ≥ 22.

Exclusion criteria: 1) Pregnancy; 2) Participation in other studies within 30 days; 3) Undergoing of ECT or repetitive transcranial magnetic stimulation (rTMS) treatment within the past 6 months; 4) Previous poor response to ECT; 5) History of substance abuse disorder; 6) Presence of contraindications to ECT or anesthesia risks; 7) History of epilepsy.

### 2.2 i-HECT procedures (Figure 1)

#### 2.2.1 Basic settings

The i-HECT protocol comprised two treatments: ECT (1.5 ST) and LCE (0.5 ST), consistent with our previous report [3]. ECT/LCE sessions (three times per week) were performed using a spECTRUM 5000Q ECT instrument (MECTA Corporation, OR, USA), with pulse width of 1 ms and a fixed current of 800 mA. The dose titration procedure was performed to determine the ST during the first session. Subsequent treatments were either ECT or LCE based on whether the patient met the criteria for ECT-LCE transition.

**Figure 1.**
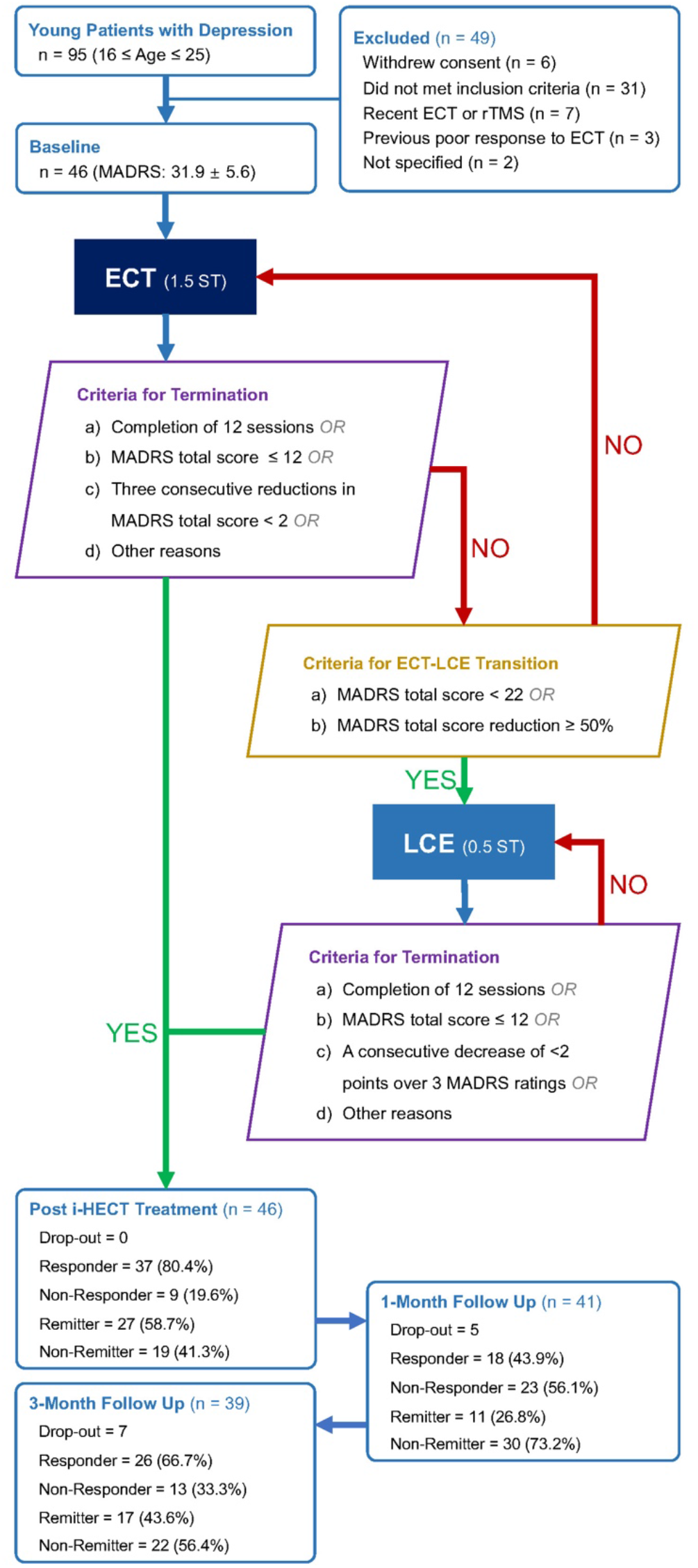
Flow chart. Abbreviations: ECT: electroconvulsive therapy; LCE: low-charge electrotherapy; ST: seizure threshold; rTMS: repetitive transcranial magnetic stimulation; MADRS: Montgomery-Åsberg Depression Rating Scale; i-HECT: individualized Hybrid-ECT.

#### 2.2.2 Criteria for ECT-LCE transition

When either of the following conditions is met, the next treatment is switched to LCE: a) MADRS total score < 22; b) MADRS total score reduction ≥ 50%.

#### 2.2.3 Criteria for terminating i-HECT treatment

After meeting any of the following criteria, i-HECT treatment was terminated and transitioned to the follow-up period: a) Completion of 12 ECT/LCE sessions; b) MADRS total score < 12; c) A consecutive decrease of < 2 points over three MADRS assessments; d) Other reasons, such as voluntary withdrawal or other medical considerations.

### 2.3 Visit Schedule

The five visits were set at 1) baseline; 2) post-ECT-LCE-transition; 3) post-treatment (within 24–48 hour after the last ECT/LCE session); 4) end of the 1-month follow-up period; and 5) end of the 3-month follow-up period.

### 2.4 Pharmacotherapy

Patients maintained their antidepressants and antipsychotics during the trial. Anticonvulsants, mood stabilizers, or lithium were discontinued during the i-HECT treatment. Single dose of alprazolam or oxazepam were prescribed as necessary when patients became agitated or felt anxious, but were prohibited 24 hour before ECT/LCE sessions. When patients suffered from insomnia, zopiclone, eszopiclone, or zolpidem were temporarily prescribed.

### 2.5 Statistical analysis

The primary outcome is the response rate analyzed in the intent-to-treat sample. Response is defined as a reduction of ≥ 50% in MADRS total score relative to baseline at the post-treatment visit. Additionally, achieving a MADRS score ≤ 12 is considered remission.

The trial employed an optimal Simon’s two-stage design [4], with a null threshold referencing O’Reardon et al.’s RCT set at 18.1% [7], and an alternative threshold referencing Kellner et al.’s RCT set at 43% [8]. In stage I, 19 patients will be recruited. If 4 or fewer responders are observed, the trial will be halted prematurely. Otherwise, an additional 27 patients will be recruited in stage II. If 13 or more responses among these total 46 patients, the null hypothesis will be rejected, indicating promise for the i-HECT treatment. The design controls the type I error rate at 0.05 and yields the power of 0.95. The secondary outcomes including MADRS and its subscales, 9-item Patient Health Questionnaire (PHQ-9) [9], Generalized Anxiety Disorder 7 (GAD-7) [10], Columbia–Suicide Severity Rating Scale (C-SSRS) [11], and Positive and Negative Syndrome Scale (PANSS) [12]. The subjective cognitive functions were evaluated using Subjective Cognitive Decline Questionnaire, and the objective cognitive functions were evaluated using Repeatable Battery for the Assessment of Neuropsychological Status (RBANS) [13], and Stroop Color and Word Test [14]. Orientation recovery tests (ORTs) after each ECT/LCE session were used to measure recovery time after ECT/LCE. Any adverse events (AEs) or drop-out for any reason was recorded to analyze the safety. The linear mixed model and the least square mean (LSM) and its 95% confidence intervals (CIs) were used to estimate the effects of outcomes at each visit using the *LmerTest* package [15] in R (version 4.2.0).

## 3. Results

A total of 46 patients were enrolled. The baseline characteristics are summarized in **Table 1**.

**Table 1.** Baseline Characteristics.

| Characteristic | Results |
| --- | --- |
| Sex, Female, n (%) | 32 (69.6%) |
| Age, year, mean ± SD | 18.7 ± 2.3 |
| Education Years, mean ± SD | 11.8 ± 1.9 |
| Diagnosis, MDD/BPD, n (%) | 27 (58.7%)/19 (41.3%) |
| With Psychotic Feature, n (%) | 19 (41.3%) |
| Onset Age, year, mean ± SD | 15.8 ± 2.9 |
| Disease Course, year, mean ± SD | 2.7 ± 1.5 |
| Current Episode, month, mean ± SD | 3.8 ± 4.3 |
| History of ECT, n (%) | 5 (10.9%) |
| Equivalent Fluoxetine Dose, mg/day, mean ± SD | 46.5 ± 16.2 |
| Equivalent Olanzapine Dose, mg/day, mean ± SD | 8.7 ± 3.8 |
| Montgomery-Åsberg Depression Rating Scale (MADRS) | 31.9 ± 5.6 |
| Total Score, mean ± SD | |
| Cognitive Pessimism, mean ± SD | 14.4 ± 2.3 |
| Affective, mean ± SD | 9.1 ± 2.3 |
| Cognitive Anxiety, mean ± SD | 5.1 ± 1.8 |
| Vegetative, mean ± SD | 6.2 ± 2.0 |
| 9-item Patient Health Questionnaire (PHQ-9), mean ± SD | 19.3 ± 5.4 |
| Generalized Anxiety Disorder 7 (GAD-7), mean ± SD | 12.1 ± 5.6 |
| Columbia–Suicide Severity Rating Scale (C-SSRS) |  |
| Suicidal Ideation and Attempts, mean ± SD | 3.5 ± 1.1 |
| Intensity of Ideation, mean ± SD | 16.3 ± 4.2 |
| Stopped Suicide Attempt, mean ± SD | 3.0 ± 1.6 |
| Positive and Negative Syndrome Scale (PANSS) |  |
| Positive Scale, mean ± SD | 13.0 ± 2.3 |
| Negative Scale, mean ± SD | 17.9 ± 5.0 |
| General Psychopathology Scale, mean ± SD | 37.1 ± 5.4 |
| Subjective Cognitive Decline Questionnaire, mean ± SD | 14.4 ± 4.3 |
| The Repeatable Battery for the Assessment of Neuropsychological Status (RBANS) |  |
| Total Score, mean ± SD | 396.1 ± 55.6 |
| Immediate Memory, mean ± SD | 70.8 ± 14.1 |
| Visuospatial/Constructional, mean ± SD | 83.4 ± 13.4 |
| Language, mean ± SD | 69.0 ± 18.5 |
| Attention, mean ± SD | 97.0 ± 14.4 |
| Delayed Memory, mean ± SD | 76.0 ± 19.6 |
| Stroop Color and Word Test |  |
| Error Interference Score, mean ± SD | 2.0 ± 1.9 |
| Time Interference Score, second, mean ± SD | 39.1 ± 11.0 |

### 3.1 Efficacy

All patients completed the treatments. Five patients did not complete the 1-month follow-up, and seven patients did not complete the 3-month follow-up (**Figure 1**). At the post-treatment visit, 37 patients (80.4%) achieved a response, with 27 (58.7%) achieving remission. The mean number of treatment sessions was 6.0 + 2.1, with 3.0 + 1.7 sessions of ECT and 3.2 + 1.6 sessions of LCE. Forty-three patients transitioned from ECT to LCE due to meeting the ECT-LCE transition criteria. Three patients did not meet the criteria and remained on ECTs throughout the trial, all three were non-responders. The reduction in MADRS score was 19.1 + 7.0, with Hedges’ *g* = 3.29 (95% CI = 2.66 to 3.91). For details, please see **sFigure 1 and Table 2**.

**Table 2.**
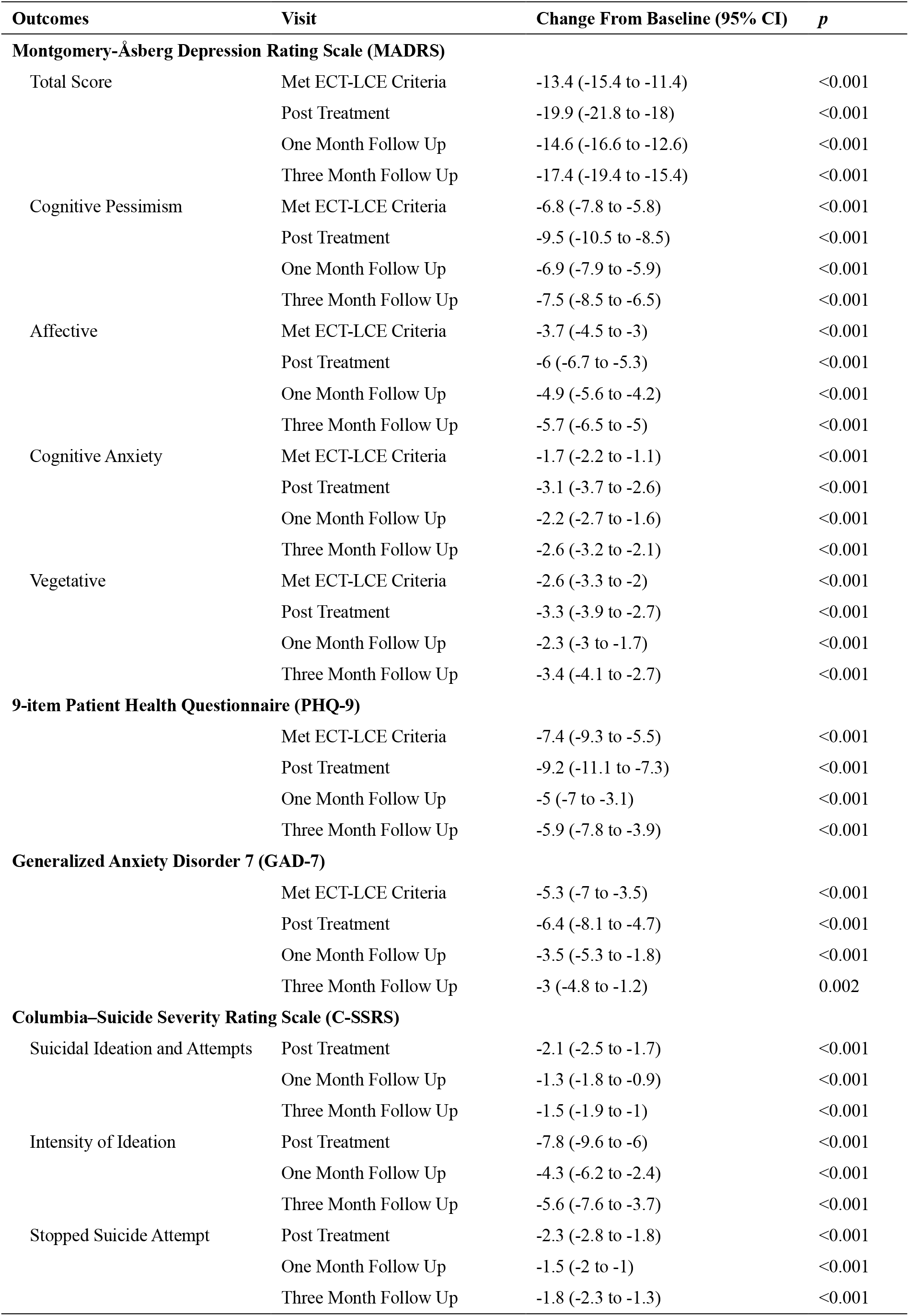
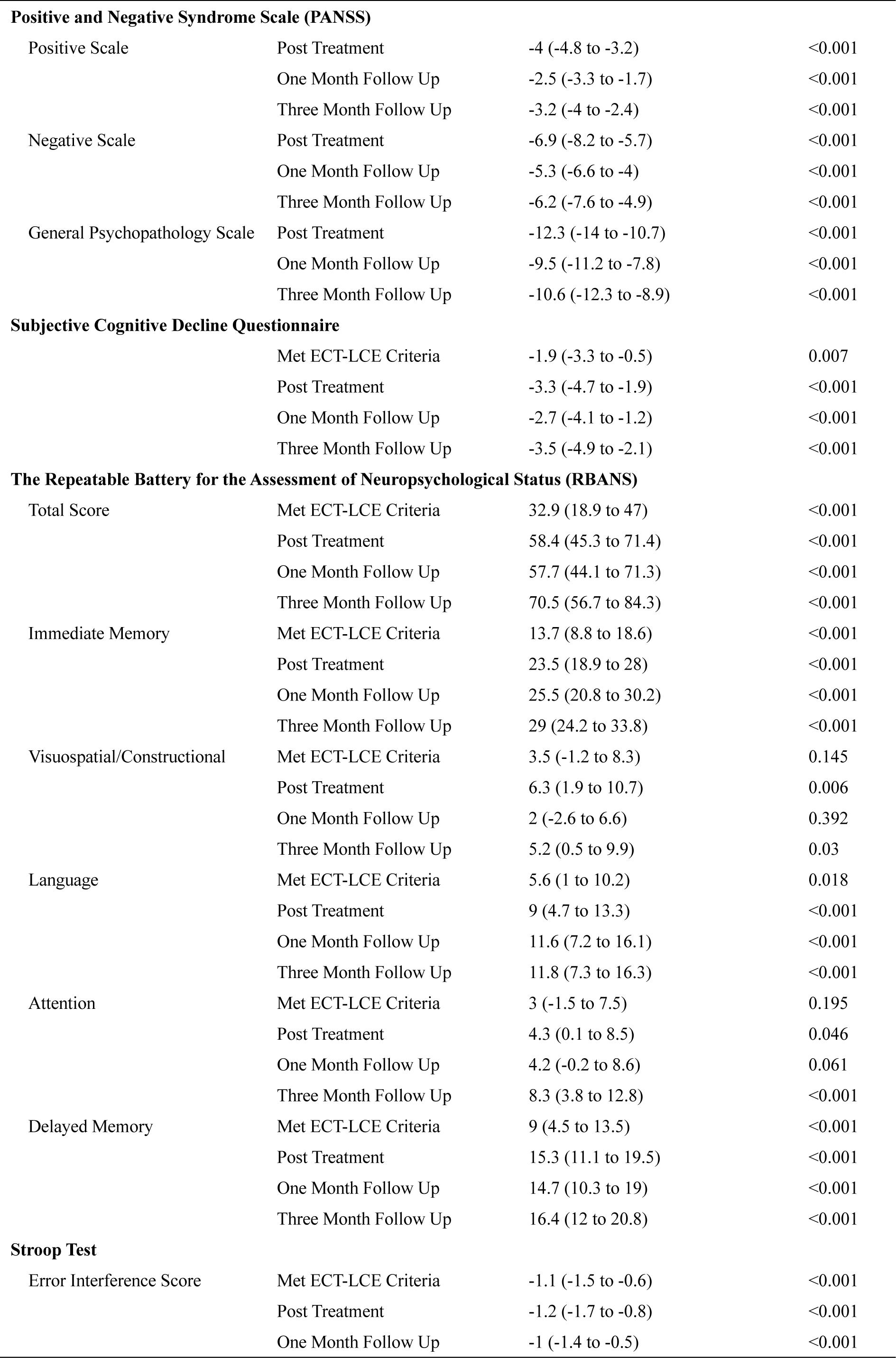
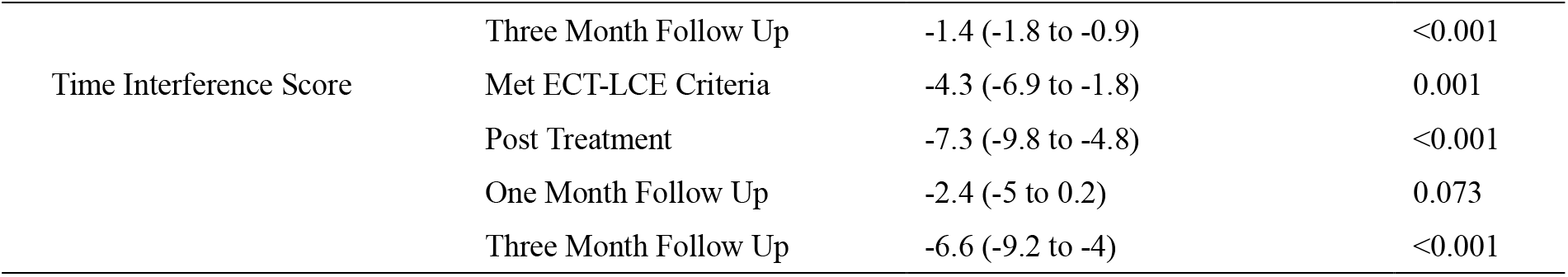
Clinical and Cognitive Outcomes.

Following i-HECT treatments, significant improvements were observed across various measures including MADRS, PHQ-9, GAD-7, C-SSRS, and PANSS. Notably, cognitive function, a key focus of this trial, demonstrated significant improvements in both subjective and objective assessments. (**sFigure 2–4**, and **Table 2**)

### 3.2 Safety

The most commonly reported AEs were memory complaints, headaches, and fatigue. The incidence of these three AEs, along with nausea and dizziness, are notably lower after LCE compared to ECT, and the ORTs after LCEs were significantly shorter than ECTs (**Table 3**).

**Table 3.**
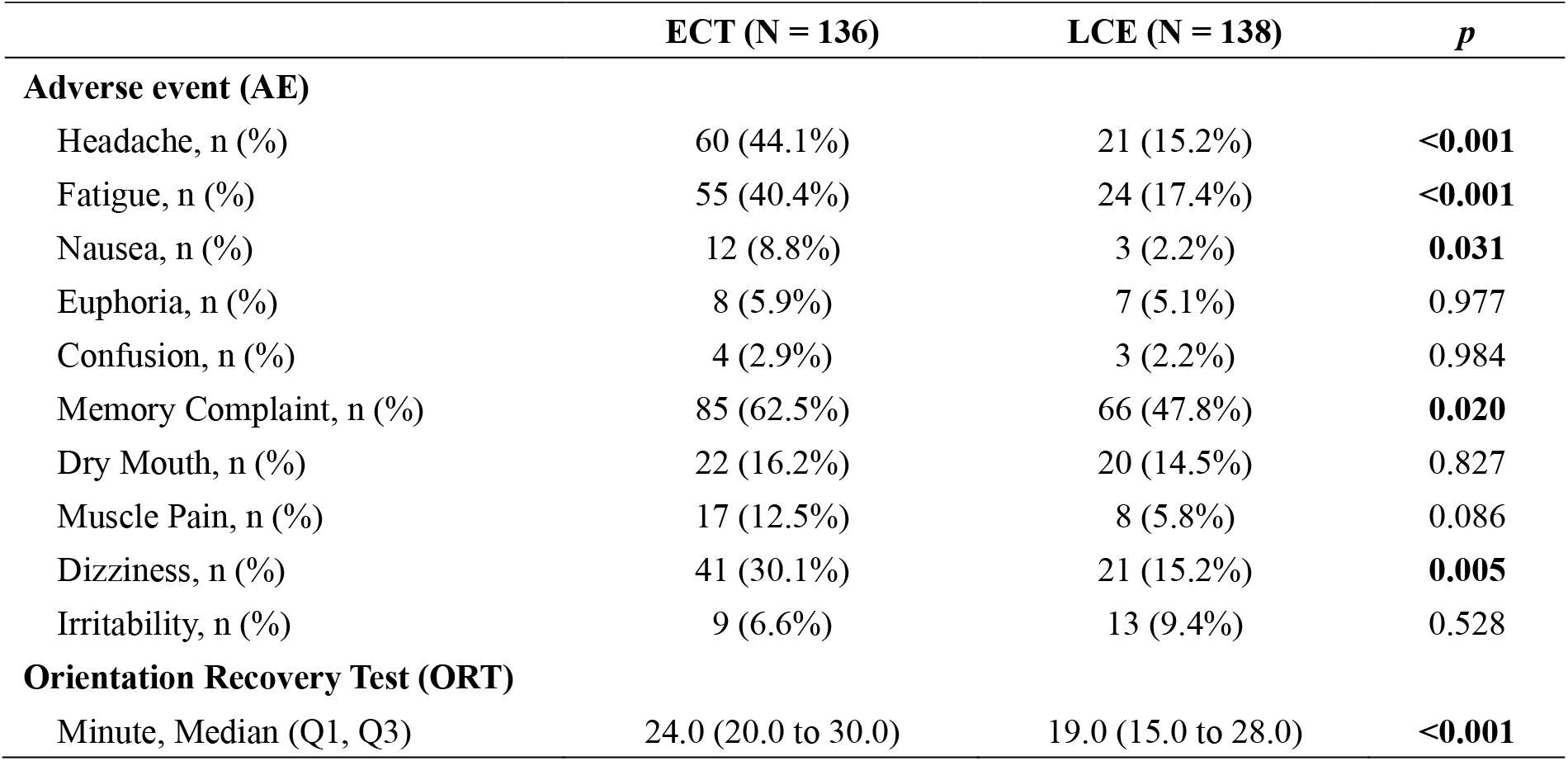
Safety Results.

### 3.3 Data accessibility

The data that support the findings of this study are available from the corresponding author, upon reasonable request.

## 4. Discussion

In brief, i-HECT showed comparable antidepressant efficacy to ECT. Moreover, the most promising outcome was the significant improvement observed in both subjective and objective cognitive functions post-treatment, with this trend continuing during the 3-month follow-up period. Young patients with depression are typically in early stages of their education or career, making prompt intervention imperative. While ECT is a most evidence-supported rapid antidepressant treatment, concerns about cognitive side effects have reduced its use in this population of young patients with depression. However, our i-HECT protocol may provide a new and simple option for addressing these concerns in these young individuals.

One major advantage of i-HECT is its compatibility with existing ECT equipment worldwide, requiring no specialized modifications. Another advantage is its simple setup, requiring no additional training for operators.

In conclusion, our i-HECT protocol offers promise as a rapid antidepressant treatment for young patients with depression, with the added benefit of improving cognitive function. Its simplicity makes it a potentially valuable treatment option.

## Supplementary Materials

**sFigure 1.**
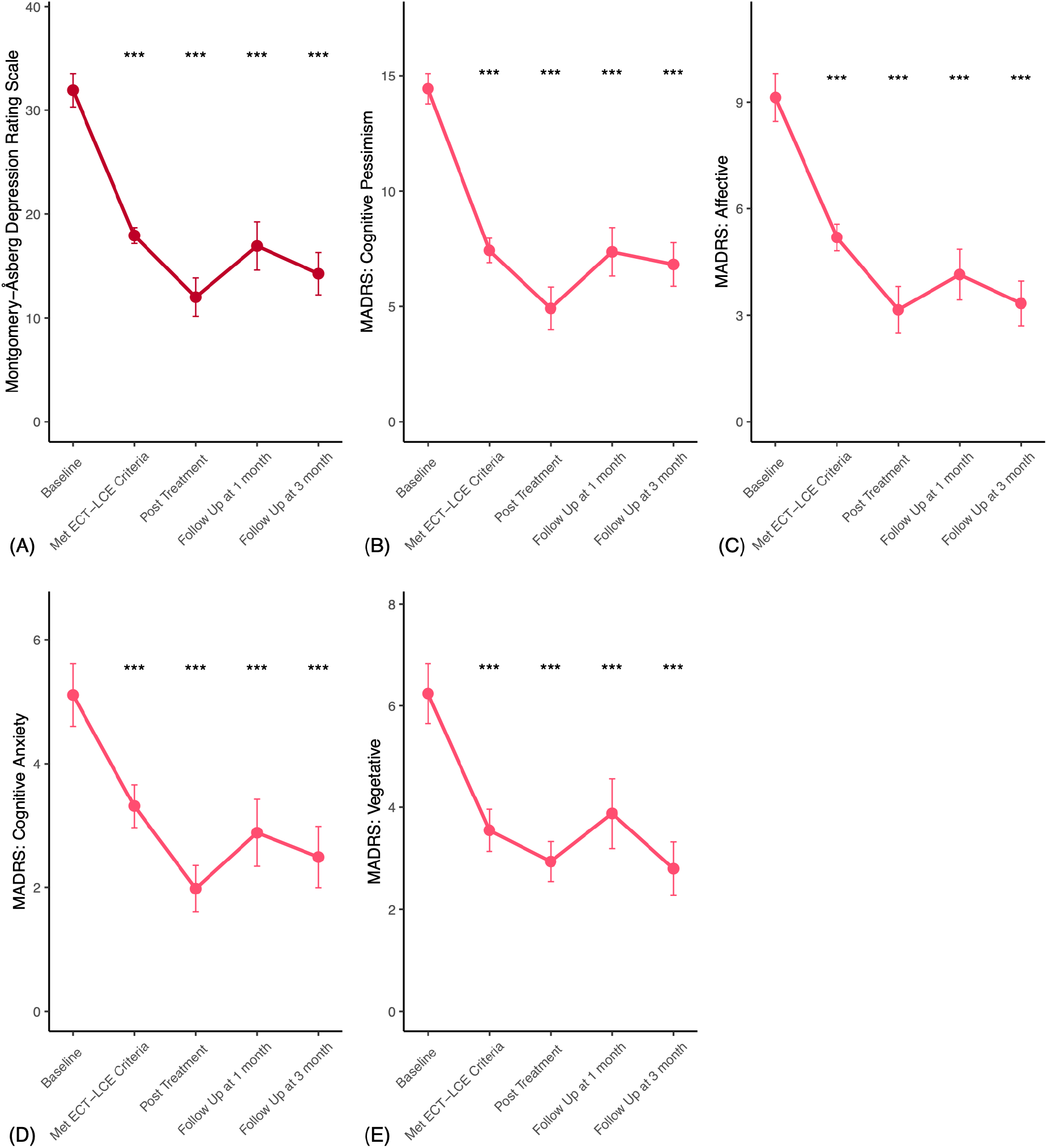
Changes in Montgomery-Åsberg Depression Rating Scale (MADRS). Note: Error bars represent 95% confidence intervals (CIs). \**p* < 0.05, \*\**p* < 0.01, \*\*\**p* < 0.001. Abbreviations: ECT: electroconvulsive therapy; LCE: low-charge electrotherapy.

**sFigure 2.**
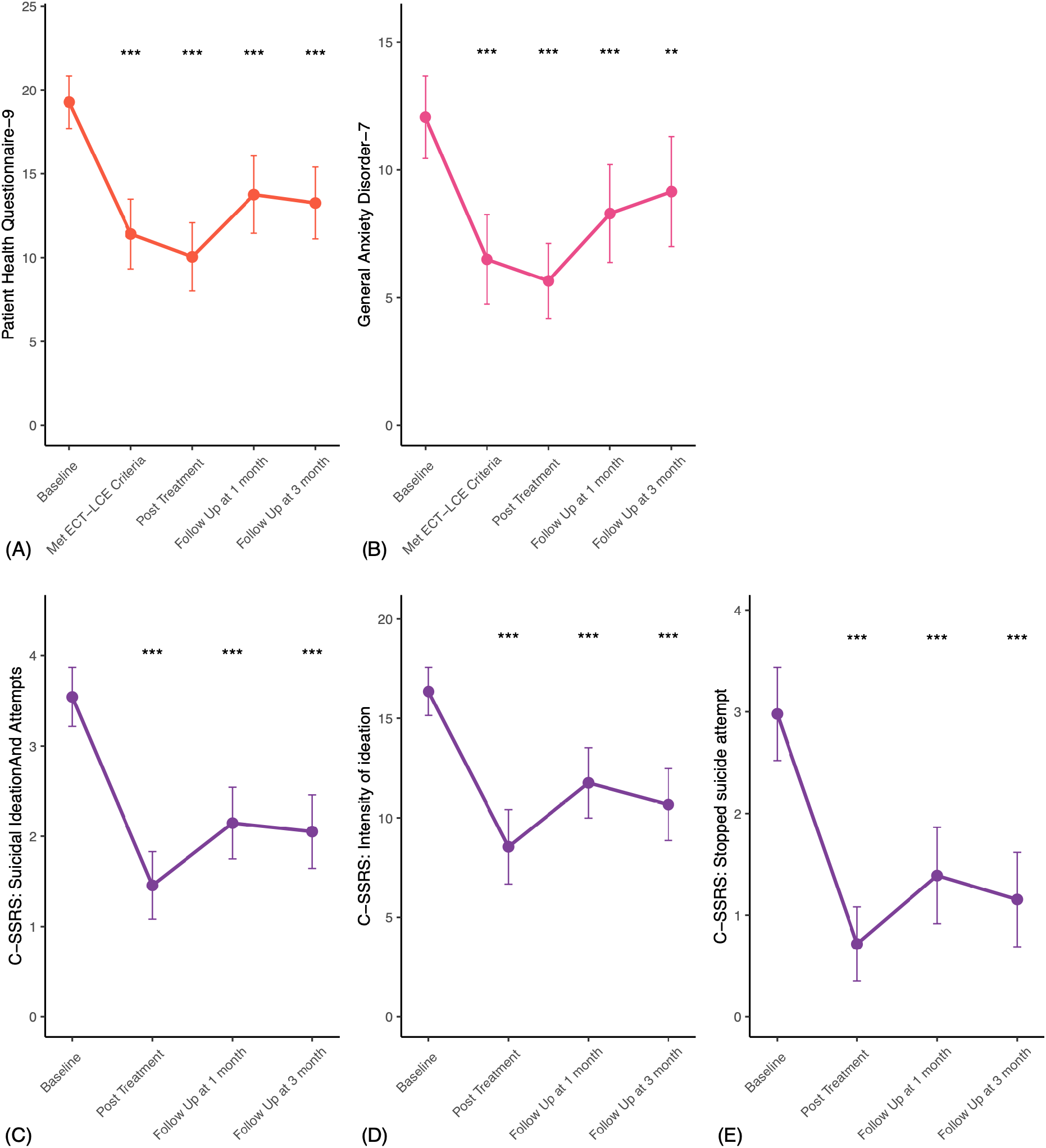
Changes in 9-item Patient Health Questionnaire (PHQ-9), Generalized Anxiety Disorder 7 (GAD-7), and Columbia–Suicide Severity Rating Scale (C-SSRS). Note: Error bars represent 95% confidence intervals (CIs). \**p* < 0.05, \*\**p* < 0.01, \*\*\**p* < 0.001. Abbreviations: ECT: electroconvulsive therapy; LCE: low-charge electrotherapy.

**sFigure 3.**
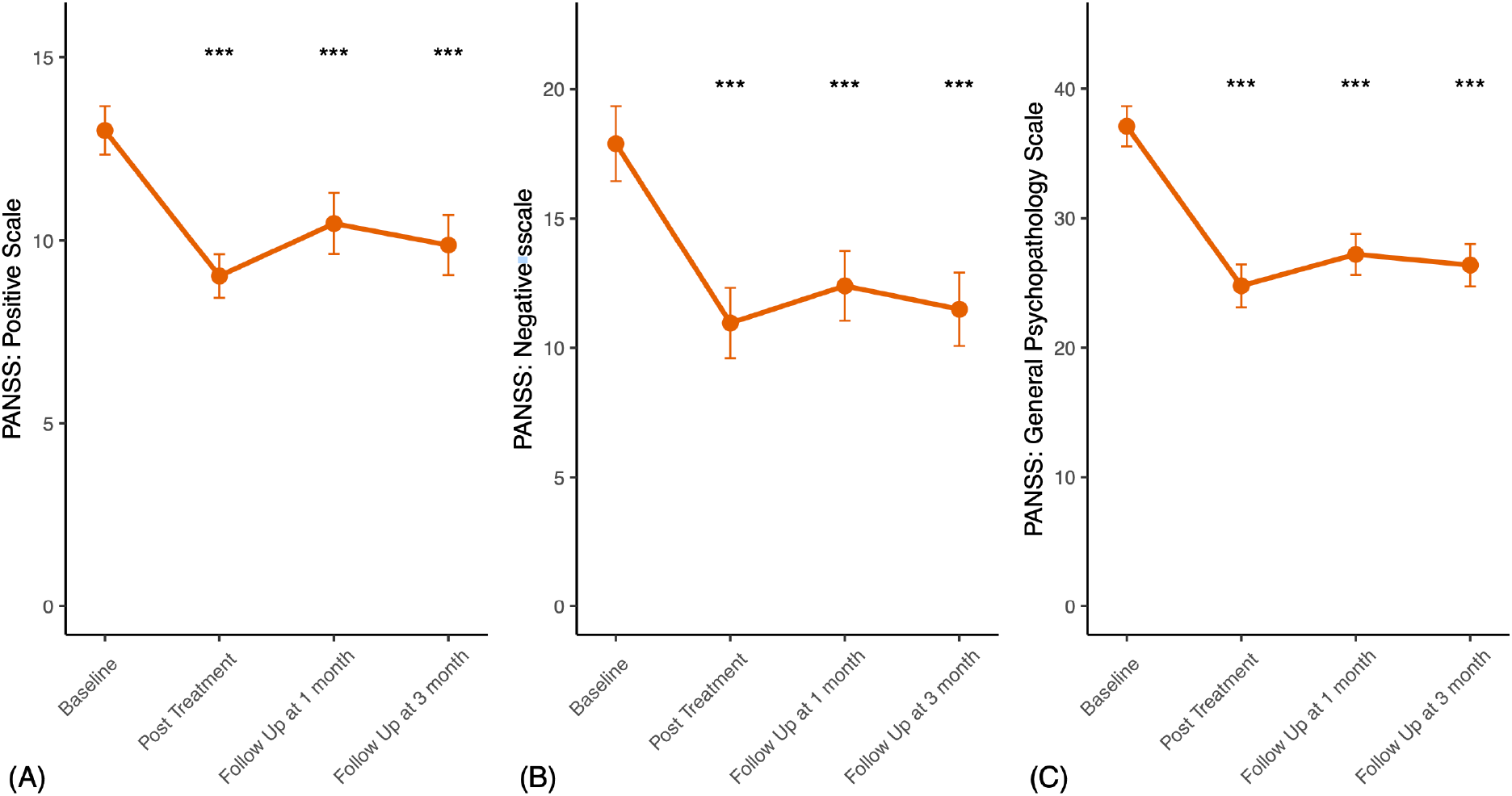
Changes in Positive and Negative Syndrome Scale (PANSS). Note: Error bars represent 95% confidence intervals (CIs). \**p* < 0.05, \*\**p* < 0.01, \*\*\**p* < 0.001. Abbreviations: ECT: electroconvulsive therapy; LCE: low-charge electrotherapy.

**sFigure 4.**
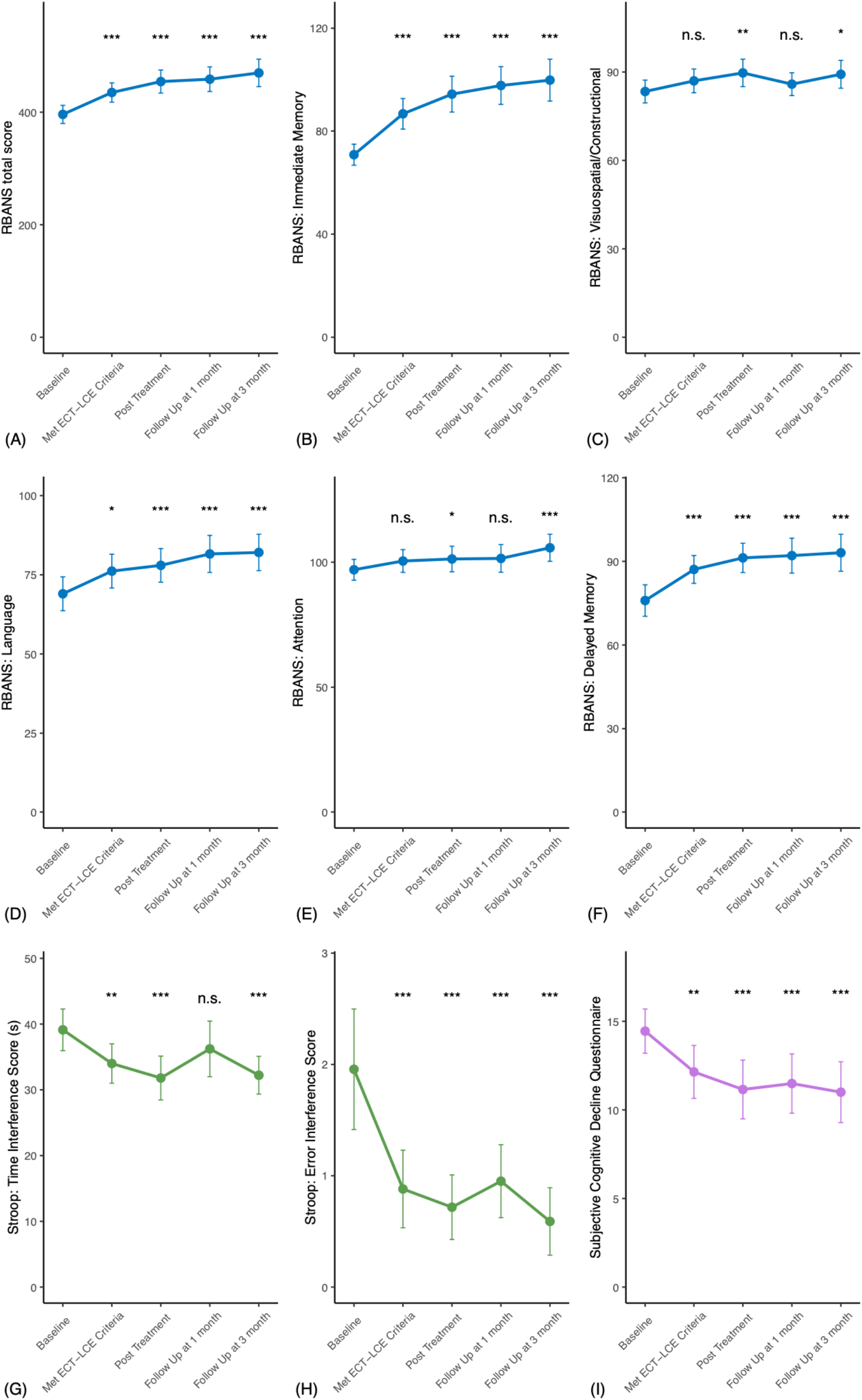
Changes in Objective (RBANS and Stroop tests) and Subjective Cognitive (SCDQ) Functions. Note: Error bars represent 95% confidence intervals (CIs). \**p* < 0.05, \*\**p* < 0.01, \*\*\**p* < 0.001. Abbreviations: RBANS: Repeatable Battery for the Assessment of Neuropsychological Status. SCDQ: Subjective Cognitive Decline Questionnaire; ECT: electroconvulsive therapy; LCE: low-charge electrotherapy.

